# TruBlk™ Shilajit Resin Supplementation Improves Muscle Strength, Endurance, and Exercise Recovery in Males Undertaking Resistance Training: A Randomised, Double-Blind, Placebo-Controlled, Multicenter Trial

**DOI:** 10.64898/2026.07.27.26358996

**Authors:** Divya Yadav, Rohit Gupta, Aashish Chaudhary, Sanjay Mishra, Santosh Ghai, Preeti Chhabra, Manoj Karwa, Surya Teja Reddy, Amandeep Singh, Karan M. Shah

## Abstract

Shilajit is a naturally occurring resinous exudate with centuries of documented use in Ayurvedic medicine. Its principal bioactive constituents — fulvic acid and a proprietary aromatic compound complex termed shilarathenes™, comprising urolithin metabolites, phenolic acids, and flavonoids — are proposed to enhance mitochondrial ATP synthesis, attenuate exercise-induced muscle damage, and modulate androgen biosynthesis. Prior controlled trials have demonstrated increases in testosterone and retention of muscular strength with purified shilajit, though large trials in resistance-trained populations have been absent.

In this randomised, double-blind, placebo-controlled, multicenter trial (CTRI/2025/07/091323), 100 healthy males aged 21–50 years (BMI <30 kg/m²; ≥1 year of resistance training) were enrolled across four sites. Participants received 250 mg twice daily of standardised TruBlk™ Shilajit resin (≥60% fulvic acid; ≥10% shilarathenes™) or matched placebo for 90 days, and were instructed to maintain habitual training and dietary practices throughout. Primary endpoints were changes from baseline in 1RM leg press, muscle endurance, RPE, and DOMS. Secondary endpoints included VO₂max, testosterone, creatine kinase, lactate dehydrogenase, handgrip strength, and global improvement ratings.

Ninety-nine participants completed the trial (99% retention). All primary outcomes showed significantly steeper improvement in the active group, assessed by treatment × time interaction: 1RM leg press (+26.0% vs +18.4%; p=0.0003, d=0.74), muscle endurance (+110.8% vs +80.5%; p=0.0017, d=0.60), RPE (p=0.002), and DOMS (p=0.036). Dominant handgrip strength showed the largest secondary effect (+12.5% vs +7.9%; p=0.0002, d=0.77). The active group also demonstrated greater reductions in creatine kinase (−33.6% vs −6.9%; p=0.030, δ=−0.26) and lactate dehydrogenase (−17.8% vs −4.2%; p=0.039, δ=−0.24), greater increases in free testosterone (+21.8% vs −2.9%; p=0.030, d=0.45) and total testosterone (+15.6% vs −3.8%; p=0.038, d=0.43), and a modest improvement in VO₂max (+3.8% vs +2.9%; p=0.032, d=0.41) compared with placebo. No adverse events were recorded in the active group.

Ninety days of TruBlk™ Shilajit resin supplementation produced robust improvements in muscle strength and endurance in resistance-trained males, with moderate effect sizes for the strength outcomes. These data support TruBlk™ Shilajit resin as a safe and promising ergogenic agent meriting further clinical evaluation.

## 1. Introduction

Shilajit (*Asphaltum punjabinum*), also known as mumie or moomiyo, is a dark tar-like resinous exudate found seeping from rock crevices in high-altitude mountain ranges, including the Himalayas, Altai, and Caucasus [1–3]. Ayurvedic medicine has valued it for centuries as a potent rejuvenator (*Rasayana*) and cognitive enhancer (*medhya rasayana*), with historical documentation of its use in fatigue, metabolic disorders, infertility, and age-related decline [4–6]. Modern research has since substantiated several of these traditional claims, reporting adaptogenic, antioxidant, anti-inflammatory, and immune-modulating properties [1, 7, 8], and generating considerable interest in Shilajit as a sports nutrition supplement [9, 10].

Shilajit’s biological activity is attributed primarily to two constituent fractions. Fulvic acid is the principal active compound [1, 11]. Its chemical structure is rich in carboxyl, hydroxyl, and phenolic groups, which together confer strong antioxidant and chelating properties alongside an ability to permeate cell membranes, facilitating nutrient transport and mitochondrial support [8]. The second fraction is a naturally occurring spectrum of dibenzo-α-pyrones (DBPs), urolithin metabolites, phenolic acids, and flavonoids, we collectively termed shilarathenes™[12]. Notably, DBP denotes a broad structural class rather than a single standardised chemical entity: the two principal markers historically quantified under this term in shilajit pharmacology, 3-hydroxydibenzo-α-pyrone and 3,8-dihydroxydibenzo-α-pyrone, are chemically identical to urolithin B and urolithin A respectively, and are considered throughout this manuscript as part of the wider shilarathenes™ bioactive spectrum [13].

These component classes each contribute distinct but complementary mechanisms. DBPs have been proposed to function as electron shuttles within the mitochondrial electron transport chain, supporting ATP production and mitochondrial biogenesis [14]. Urolithin metabolites support mitochondrial quality control through selective mitophagy, clearing damaged mitochondria and sustaining energy output during prolonged exercise [15]. Phenolic acids reduce exercise-induced oxidative damage and suppress inflammatory signalling through the NF-κB pathway [16, 17], whilst flavonoids stimulate nitric oxide production through eNOS activation, broadening blood vessel diameter and improving oxygen delivery to working muscle [18]. Beyond energy metabolism, these constituents are also proposed to support testosterone biosynthesis through direct actions at the level of the testes, upregulating steroidogenic enzyme activity, inhibiting aromatase, and improving Leydig cell responsiveness to gonadotrophic stimulation, without disrupting the hypothalamic-pituitary axis [10, 19].

A persistent challenge in Shilajit research has been the considerable variability in bioactive content and purity across commercial preparations. TruBlk™, the extract used in this trial, addresses this through HPLC-validated standardisation of both fulvic acid (≥60% by weight) and shilarathenes™ (≥10% by weight), providing a reproducible and authenticated bioactive profile that distinguishes it from unstandardised commercial resins.

Prior clinical evidence, whilst encouraging, has been limited in scale and scope. Pandit et al. demonstrated significant increases in free and total testosterone with purified Shilajit at 250 mg twice daily over 90 days in healthy males aged 45 to 55 years. Keller et al. (2019) reported dose-dependent preservation of maximal muscular strength alongside reductions in serum hydroxyproline, a marker of connective tissue degradation, following eight weeks of supplementation in recreationally active men, though these effects were observed only in the higher dose group [9]. Das et al. (2016) observed upregulation of skeletal muscle repair and collagen remodelling genes following Shilajit supplementation, though the absence of a structured exercise stimulus in that cohort may account for the lack of corresponding changes in muscle damage markers [20]. A recent open-label pilot study with the TruBlk™ formulation offered more direct preliminary support, reporting significant improvements in handgrip strength, VO₂max, and muscle damage markers over 28 days in healthy adults, with no adverse events [21]. Taken together, these findings point to a biologically plausible and clinically promising ergogenic profile, yet no large, adequately powered, multicenter, placebo-controlled trial has examined the effects of standardised Shilajit resin on exercise performance, hormonal, and recovery outcomes in resistance-trained males, a population for whom evidence-based ergogenic support is of particular practical relevance.

Resistance training places substantial and repeated demands on the muscular, metabolic, and endocrine systems, inducing oxidative stress, inflammation, and fatigue that can progressively limit performance and recovery between sessions [22, 23]. Safe and effective ergogenic aids that support training adaptation are therefore of considerable interest to athletes and sports medicine practitioners [24]. Given Shilajit’s proposed mechanisms, its demonstrated effects on testosterone and muscle preservation in prior trials, and the preliminary signal from the TruBlk™ pilot study, we hypothesised that supplementation with standardised Shilajit resin would produce meaningful improvements in strength, endurance, recovery, and hormonal profile in resistance-trained males. This trial was designed to test that hypothesis. We report the findings of a randomised, double-blind, placebo-controlled, multicenter trial examining the efficacy and safety of TruBlk™ Shilajit resin at 250 mg twice daily over 90 days, evaluated across a comprehensive range of performance, recovery, and hormonal outcomes.

## 2. Methods

### 2.1 Study Design

This was a randomised, double-blind, placebo-controlled, parallel-group, multicenter trial conducted across four investigational sites between 30th August 2025 and 6th January 2026 (CTRI/2025/07/091323). The trial was conducted in accordance with ICH-GCP guidelines and the Declaration of Helsinki, with institutional review board approval obtained at all four sites prior to commencement. All participants provided written informed consent before enrolment. This manuscript was prepared in accordance with the CONSORT 2010 guidelines for reporting randomised trials [25].

### 2.2 Participants

One hundred healthy males aged 21 to 50 years were enrolled. Eligible participants had a BMI below 30 kg/m² and a minimum of one year of structured resistance training, defined as at least three sessions per week. Participants were required to maintain their habitual diet, training, and lifestyle throughout the study period and to be capable of providing written informed consent.

Individuals were excluded if they had used anabolic steroids, testosterone therapy, or hormonal supplements within the preceding six months, or other performance supplements including creatine or beta-alanine within the preceding four weeks. Further exclusion criteria included chronic medical conditions such as cardiovascular disease, diabetes, or renal and hepatic impairment; musculoskeletal injuries affecting exercise performance; known allergy to Shilajit or related compounds; and participation in another clinical trial within the preceding 30 days.

### 2.3 Randomisation and Blinding

Participants were assigned in a 1:1 ratio to either TruBlk™ or placebo using a computer-generated randomisation schedule with permuted blocks of four, stratified by site. Allocation codes were sealed in opaque envelopes and held by an independent data management team. Participants, investigators, outcome assessors, and data analysts remained blinded to treatment assignment throughout the trial.

### 2.4 Intervention

Participants in the active group received TruBlk™ Shilajit resin, a purified extract standardised to ≥60% fulvic acid and ≥10% shilarathenes™ by HPLC-validated methodology. The intervention was supplied in single-dose sachets of 250 mg, with participants instructed to take one sachet in the morning and one in the evening, both with meals, for 90 days, giving a total daily dose of 500 mg. Placebo sachets contained microcrystalline cellulose and were identical in appearance, packaging, taste, and odour to the active treatment.

Compliance was assessed by sachet count at each scheduled study visit. Participants returning fewer than 80% of expected empty sachets were flagged for per-protocol exclusion.

### 2.5 Outcome Measures

Primary and secondary endpoints were assessed at baseline, Day 30, Day 60, and Day 90.

The four primary endpoints were: one-repetition maximum (1RM) leg press, assessed as the maximum weight lifted for a single complete repetition on a 45° leg press machine using a standardised warm-up and testing protocol; muscle endurance, defined as the number of repetitions completed at 70% of 1RM to volitional fatigue under standardised conditions; rating of perceived exertion (RPE), recorded immediately following the endurance test using a standardised scale; and delayed-onset muscle soreness (DOMS), rated 48 hours post-exercise on a 10 cm visual analogue scale (0 = no soreness, 10 = extreme soreness).

Secondary endpoints comprised maximal oxygen uptake (VO₂max), estimated via the Harvard step test; fasting serum free testosterone (pg/mL) and total testosterone (ng/dL), measured by ELISA from morning blood samples collected between 07:00 and 09:00; serum creatine kinase (CK, U/L) and lactate dehydrogenase (LDH, U/L), measured 48 hours post-exercise by standard enzymatic assay; dominant and non-dominant handgrip strength (kg), assessed as the best of three trials per hand using a calibrated dynamometer; and participant and investigator global assessments of overall improvement rated on a five-point Likert scale at Day 90.

Safety endpoints included adverse events, serious adverse events, vital signs, and anthropometric measurements recorded at each visit.

### 2.6 Statistical Analysis

A sample size of 100 participants (50 per group) was calculated to provide 90% power to detect a 10% between-group difference in 1RM leg press at a two-sided significance level of α=0.05, assuming a standard deviation of 12% and a 5% dropout rate.

Three analysis populations were pre-specified: the intention-to-treat (ITT) population comprising all randomised participants; the per-protocol (PP) population comprising participants completing at least 80% of study visits with at least 80% supplement compliance; and the safety population comprising all participants receiving at least one dose.

Endpoints assessed at four timepoints (1RM leg press, muscle endurance, RPE, DOMS and VO₂max) were analysed using a mixed-effects model (REML) with treatment as a between-subjects factor and time as a within-subjects factor, without assuming sphericity (Geisser–Greenhouse correction); the treatment × time interaction was the primary test of differential response between groups. Endpoints assessed at two timepoints (handgrip strength, testosterone, and muscle damage markers) were compared by between-group analysis of change from baseline. Normality of change scores was assessed by Shapiro–Wilk test: normally distributed variables were compared by unpaired t-test and are presented as mean ± SD, with Cohen’s d as the effect size; non-normally distributed variables were compared by Mann-Whitney U test and are presented as medians with interquartile ranges, with Cliff’s delta as the effect size. Within-group changes were assessed by paired t-test or Wilcoxon signed-rank test as appropriate. Analyses were performed using GraphPad Prism version 10, except for Cliff’s delta, which was computed externally. A two-sided p-value below 0.05 was considered statistically significant.

## 3. Results

### 3.1 Participant Flow and Baseline Characteristics

One hundred and twenty participants were screened for eligibility, of whom 20 did not meet inclusion criteria and were excluded prior to randomisation. The remaining 100 were randomised equally to TruBlk™ (n=50) or placebo (n=50). All 100 participants received their allocated treatment and completed assessments at Day 30 and Day 60. One participant in the TruBlk™ group withdrew consent prior to Day 90, giving a trial completion rate of 99%. The intention-to-treat population comprised 49 TruBlk™ and 50 placebo participants; the per-protocol population was identical. No protocol deviations related to supplement compliance were recorded. Participant flow is summarised in Figure 1.

**Figure 1.**
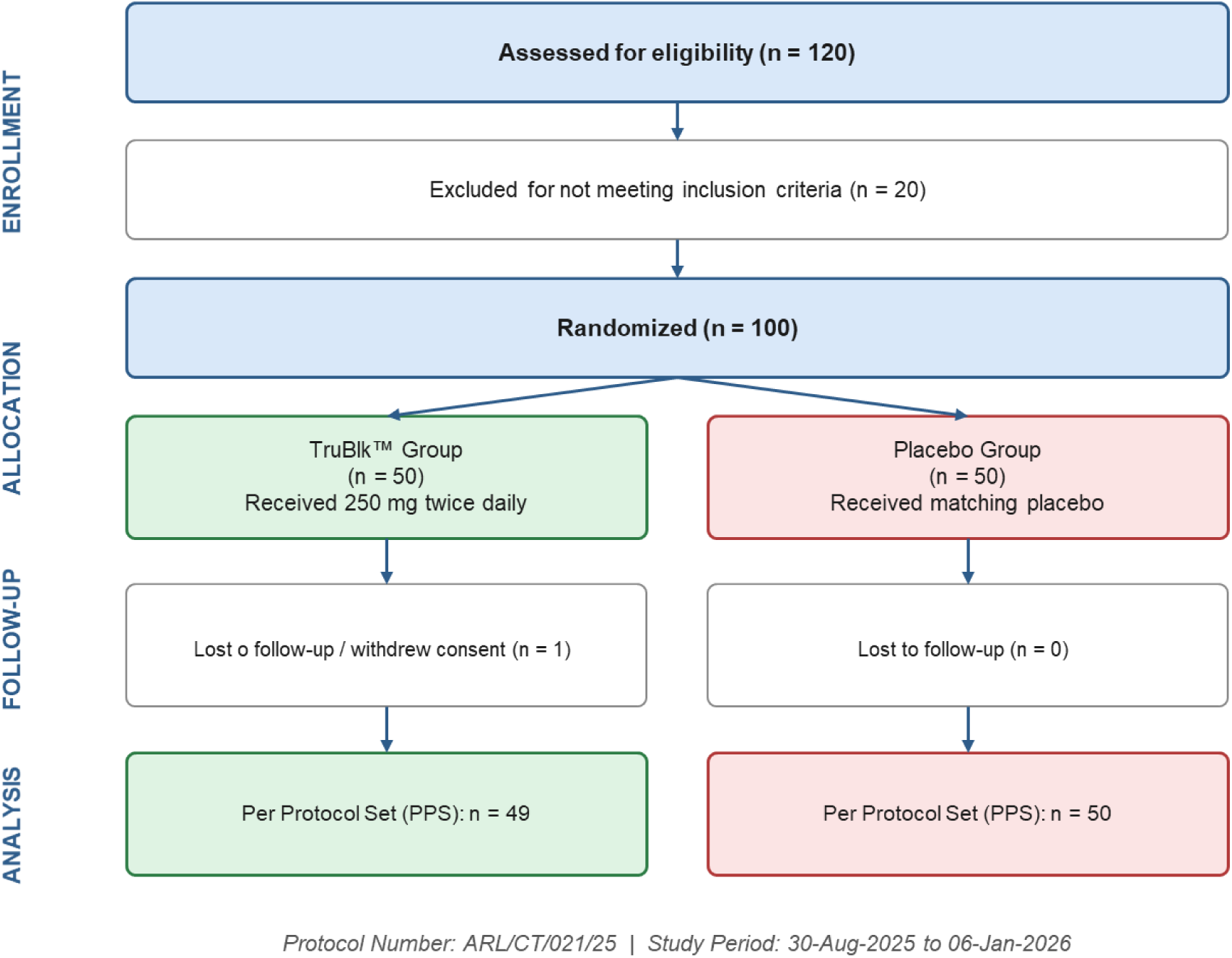
CONSORT flow diagram of subject disposition. Flow of participants through each stage of the randomized, placebo-controlled trial of TruBlk™ (CTRI/2025/07/091323). Of 120 individuals assessed for eligibility, 20 were excluded for not meeting inclusion criteria, yielding 100 participants who were randomized 1:1 to receive TruBlk™ 250 mg twice daily (n = 50) or matching placebo (n = 50). One participant in the TruBlk™ group was lost to follow-up (withdrew consent); no discontinuations occurred in the placebo group.

Groups were well matched for demographic and training characteristics at baseline. Mean age was 29.3 years in the TruBlk™ group and 28.9 years in the placebo group, with mean BMI of 24.7 and 24.5 kg/m² respectively. Average resistance training experience was 4.2 years in the TruBlk™ group and 4.0 years in the placebo group. Baseline values for all primary outcome measures were comparable between groups (all p>0.05), as were VO₂max, dominant handgrip strength, total testosterone, and CRP. Baseline CRP was 2.0 mg/L (IQR 1.2–3.4) in the TruBlk™ group and 2.2 mg/L (IQR 1.0–3.4) in the placebo group (p=0.79). Three secondary variables differed at baseline: free testosterone was lower in the TruBlk™ group (9.05 ± 3.36 vs 10.91 ± 4.17 pg/mL; p=0.02), while creatine kinase (226.2 vs 161.6 U/L; p=0.01) and lactate dehydrogenase (281.8 vs 249.3 U/L; p=0.01) were both higher. Baseline characteristics are presented in Table 1.

**Table 1.** Baseline characteristics of participants.

| Characteristic | TruBlk™ (n=49) | Placebo (n=50) | p-value |
| --- | --- | --- | --- |
| Age (years) | $29.3 \pm 6.8$ | $28.9 \pm 7.1$ | 0.78 |
| BMI (kg/m<sup>2</sup>) | $24.7 \pm 2.8$ | $24.5 \pm 2.6$ | 0.71 |
| Training experience (years) | $4.2 \pm 2.1$ | $4.0 \pm 1.9$ | 0.63 |
| **Primary outcomes** |  |  |  |
| 1RM leg press (kg) | $164.9 \pm 43.6$ | $165.0 \pm 36.8$ | 0.99 |
| Muscle endurance (reps) | $9.27 \pm 4.53$ | $9.64 \pm 4.75$ | 0.69 |
| RPE (0–10) | $6.12 \pm 1.32$ | $5.90 \pm 1.37$ | 0.46 |
| DOMS VAS (0–10) | $6.22 \pm 1.07$ | $6.12 \pm 1.21$ | 0.65 |
| **Secondary outcomes** |  |  |  |
| VO<sub>2</sub>max (mL/kg/min) | $41.49 \pm 1.19$ | $41.75 \pm 1.40$ | 0.32 |
| Dominant handgrip (kg) | $47.51 \pm 6.86$ | $47.70 \pm 7.84$ | 0.90 |
| Total testosterone (ng/dL) | $390.7 \pm 177.5$ | $405.9 \pm 146.2$ | 0.65 |
| Free testosterone (pg/mL) | $9.05 \pm 3.36$ | $10.91 \pm 4.17$ | 0.02 |
| CRP (mg/L)<sup>a</sup> | 2.0 (1.2–3.4) | 2.2 (1.0–3.4) | 0.79 |
| CK (U/L)<sup>a</sup> | 226.2 (148.1–339.0) | 161.6 (124.9–245.1) | 0.01 |
| LDH (U/L) | 281.8 (237.6–317.2) | 249.3 (214.2–274.0) | 0.01 |

Data are mean ± SD, or median (IQR) where indicated. p-values are for between-group comparison at baseline (unpaired t-test, or Mann-Whitney U test for non-normally distributed variables).
<sup>a</sup>Median (IQR); non-normally distributed.

### 3.2 Primary Endpoints

All four primary endpoints — 1RM leg press, muscle endurance, rating of perceived exertion, and delayed-onset muscle soreness — improved progressively from baseline in both groups, with significantly steeper improvement in the TruBlk™ group over the 90-day period. The results are summarised in Table 2.

**Table 2.**
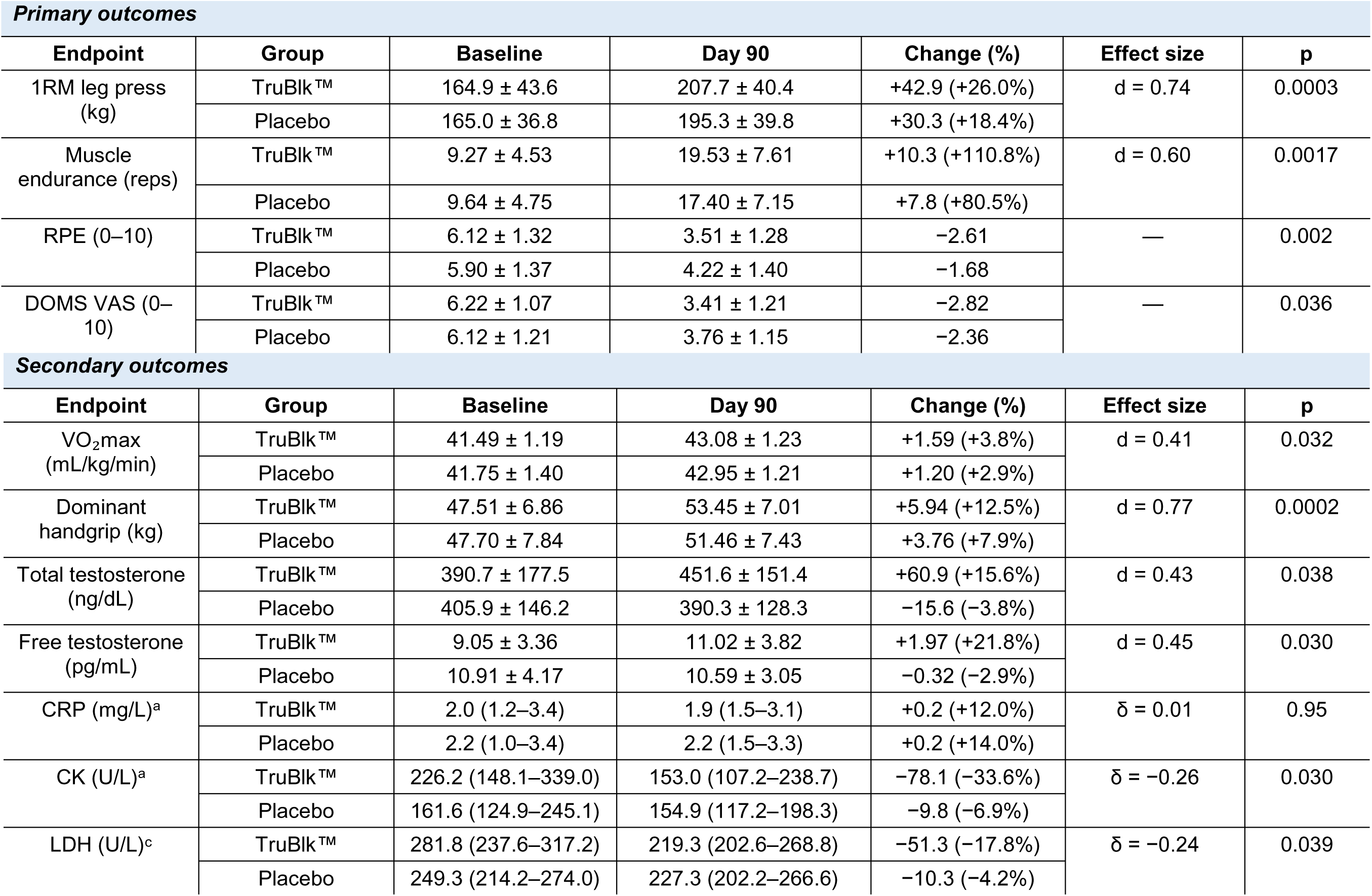
Outcome measures at baseline and day 90.

| Primary outcomes |  |  |  |  |  |  |
| --- | --- | --- | --- | --- | --- | --- |
| Endpoint | Group | Baseline | Day 90 | Change (%) | Effect size | p |
| 1RM leg press (kg) | TruBlk™ | 164.9 ± 43.6 | 207.7 ± 40.4 | +42.9 (+26.0%) | d = 0.74 | 0.0003 |
|  | Placebo | 165.0 ± 36.8 | 195.3 ± 39.8 | +30.3 (+18.4%) |  |  |
| Muscle endurance (reps) | TruBlk™ | 9.27 ± 4.53 | 19.53 ± 7.61 | +10.3 (+110.8%) | d = 0.60 | 0.0017 |
|  | Placebo | 9.64 ± 4.75 | 17.40 ± 7.15 | +7.8 (+80.5%) |  |  |
| RPE (0–10) | TruBlk™ | 6.12 ± 1.32 | 3.51 ± 1.28 | −2.61 | — | 0.002 |
|  | Placebo | 5.90 ± 1.37 | 4.22 ± 1.40 | −1.68 |  |  |
| DOMS VAS (0–10) | TruBlk™ | 6.22 ± 1.07 | 3.41 ± 1.21 | −2.82 | — | 0.036 |
|  | Placebo | 6.12 ± 1.21 | 3.76 ± 1.15 | −2.36 |  |  |
| Secondary outcomes |  |  |  |  |  |  |
| Endpoint | Group | Baseline | Day 90 | Change (%) | Effect size | p |
| VO<sub>2</sub>max (mL/kg/min) | TruBlk™ | 41.49 ± 1.19 | 43.08 ± 1.23 | +1.59 (+3.8%) | d = 0.41 | 0.032 |
|  | Placebo | 41.75 ± 1.40 | 42.95 ± 1.21 | +1.20 (+2.9%) |  |  |
| Dominant handgrip (kg) | TruBlk™ | 47.51 ± 6.86 | 53.45 ± 7.01 | +5.94 (+12.5%) | d = 0.77 | 0.0002 |
|  | Placebo | 47.70 ± 7.84 | 51.46 ± 7.43 | +3.76 (+7.9%) |  |  |
| Total testosterone (ng/dL) | TruBlk™ | 390.7 ± 177.5 | 451.6 ± 151.4 | +60.9 (+15.6%) | d = 0.43 | 0.038 |
|  | Placebo | 405.9 ± 146.2 | 390.3 ± 128.3 | −15.6 (−3.8%) |  |  |
| Free testosterone (pg/mL) | TruBlk™ | 9.05 ± 3.36 | 11.02 ± 3.82 | +1.97 (+21.8%) | d = 0.45 | 0.030 |
|  | Placebo | 10.91 ± 4.17 | 10.59 ± 3.05 | −0.32 (−2.9%) |  |  |
| CRP (mg/L)<sup>a</sup> | TruBlk™ | 2.0 (1.2–3.4) | 1.9 (1.5–3.1) | +0.2 (+12.0%) | δ = 0.01 | 0.95 |
|  | Placebo | 2.2 (1.0–3.4) | 2.2 (1.5–3.3) | +0.2 (+14.0%) |  |  |
| CK (U/L)<sup>a</sup> | TruBlk™ | 226.2 (148.1–339.0) | 153.0 (107.2–238.7) | −78.1 (−33.6%) | δ = −0.26 | 0.030 |
|  | Placebo | 161.6 (124.9–245.1) | 154.9 (117.2–198.3) | −9.8 (−6.9%) |  |  |
| LDH (U/L)<sup>c</sup> | TruBlk™ | 281.8 (237.6–317.2) | 219.3 (202.6–268.8) | −51.3 (−17.8%) | δ = −0.24 | 0.039 |
|  | Placebo | 249.3 (214.2–274.0) | 227.3 (202.2–266.6) | −10.3 (−4.2%) |  |  |

#### 1RM Leg Press

At baseline, groups were well matched for 1RM leg press, with no difference in maximal strength between those receiving TruBlk™ (164.9 ± 43.6 kg) and placebo (165.0 ± 36.8 kg; p = 0.99). Over the 90-day training period, maximal strength increased significantly in both groups (main effect of time: F(1.70, 164.6) = 341.6, p < 0.0001), consistent with the expected response to resistance training. However, the rate at which strength was gained differed between groups, with a steeper trajectory in the TruBlk™ group producing a significant treatment × time interaction (F(1.70, 164.6) = 9.57, p = 0.0003), indicating that the two groups diverged rather than improving in parallel. By day 90, participants receiving TruBlk™ had gained 42.9 kg (+26.0%) compared with 30.3 kg (+18.4%) in the placebo group, a between-group difference of 12.6 kg (95% CI 5.8 to 19.3) that represented a moderate effect (Cohen’s d = 0.74, p = 0.0004) (Figure 2A).

**Figure 2.**
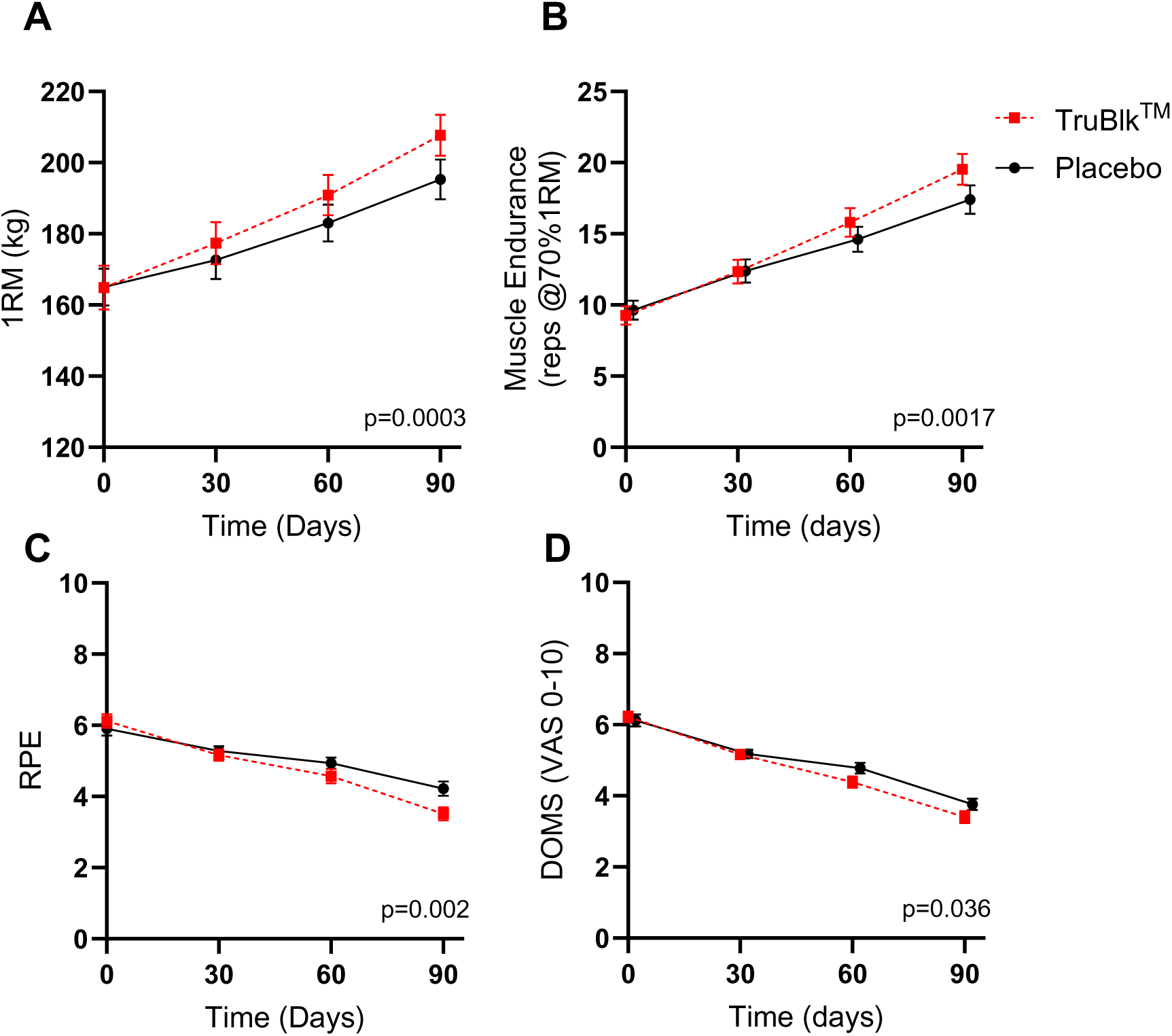
Strength, endurance and perceptual outcomes over 90 days of resistance training. (A) One-repetition maximum (1RM) leg press, (B) muscle endurance (repetitions completed at 70% of 1RM), (C) ratings of perceived exertion (RPE) and (D) delayed-onset muscle soreness (DOMS), assessed at baseline and at days 30, 60 and 90 in participants receiving TruBlk™ (red, n = 49) or placebo (black, n = 50). RPE and DOMS were rated on a 0–10 visual analogue scale. Data are mean ± SEM. P values in each graph denotes the treatment × time interaction.

#### Muscle Endurance (ME)

A comparable effect was observed for muscle endurance, with groups again well matched at baseline for the number of repetitions completed at 70% of 1RM (TruBlk™ 9.27 ± 4.53 reps; placebo 9.64 ± 4.75 reps; p = 0.69). As with maximal strength, endurance improved significantly in both groups over the 90-day period (main effect of time: F(1.45, 140.4) = 366.7, p < 0.0001), but the rate of improvement again differed between arms, with a steeper trajectory in the TruBlk™ group producing a significant treatment × time interaction (F(1.45, 140.4) = 8.13, p = 0.0017). By day 90, participants receiving TruBlk™ had gained 10.3 repetitions (+110.8%) compared with 7.8 repetitions (+80.5%) in the placebo group, a between-group difference of 2.5 repetitions (95% CI 0.8 to 4.2) that likewise represented a moderate effect (Cohen’s d = 0.60, p = 0.004) (Figure 2B).

#### Rating of Perceived Exertion (RPE)

The perceptual outcomes followed the same pattern. Groups were well matched at baseline for rating of perceived exertion (TruBlk™ 6.12 ± 1.32; placebo 5.90 ± 1.37; p = 0.46), and RPE decreased significantly over the 90-day period in both groups (main effect of time: F(2.26, 219.6) = 123.3, p < 0.0001), indicating that perceived exertion improved with continued training irrespective of supplementation. As with the performance measures, however, the improvement was greater in the TruBlk™ group, producing a significant treatment × time interaction (F(2.26, 219.6) = 5.95, p = 0.002). By day 90, RPE had fallen by 2.61 points in the TruBlk™ group compared with 1.68 points in the placebo group. (Figure 2C).

#### Delayed-Onset Muscle Soreness

A similar effect was seen for muscle soreness, with groups again well matched at baseline (TruBlk™ 6.22 ± 1.07; placebo 6.12 ± 1.21; p = 0.65). DOMS decreased significantly over the 90-day period in both groups (main effect of time: F(2.40, 233.2) = 244.1, p < 0.0001), improving with continued training in both arms. Consistent with the preceding outcomes, the decline was steeper in the TruBlk™ group, producing a significant treatment × time interaction (F(2.40, 233.2) = 3.14, p = 0.036). By day 90, soreness had fallen by 2.82 points in the TruBlk™ group compared with 2.36 points in the placebo group. (Figure 2D).

### 3.3 Secondary Endpoints

Most pre-specified secondary endpoints favoured TruBlk™ at Day 90. C-reactive protein was the exception, showing no change in either group and no between-group difference (p=0.95). Given the number of secondary comparisons performed and the absence of adjustment for multiplicity, these findings are considered exploratory and should be interpreted accordingly.

#### VO₂max

VO₂max was well matched between groups at baseline (TruBlk™ 41.49 ± 1.19; placebo 41.75 ± 1.40 mL/kg/min; p = 0.32) and improved significantly over the 90-day period in both arms (main effect of time: F(2.23, 216.2) = 85.7, p < 0.0001). A significant treatment × time interaction was observed (F(2.23, 216.2) = 3.20, p = 0.032), with a marginally steeper improvement in the TruBlk™ group. By day 90, VO₂max had increased by 1.59 mL/kg/min (+3.8%) in the TruBlk™ group compared with 1.20 mL/kg/min (+2.9%) in placebo, a between-group difference of 0.40 mL/kg/min (95% CI 0.01 to 0.78; Cohen’s d = 0.41, p = 0.032) representing a small effect (Figure 3A).

**Figure 3.**
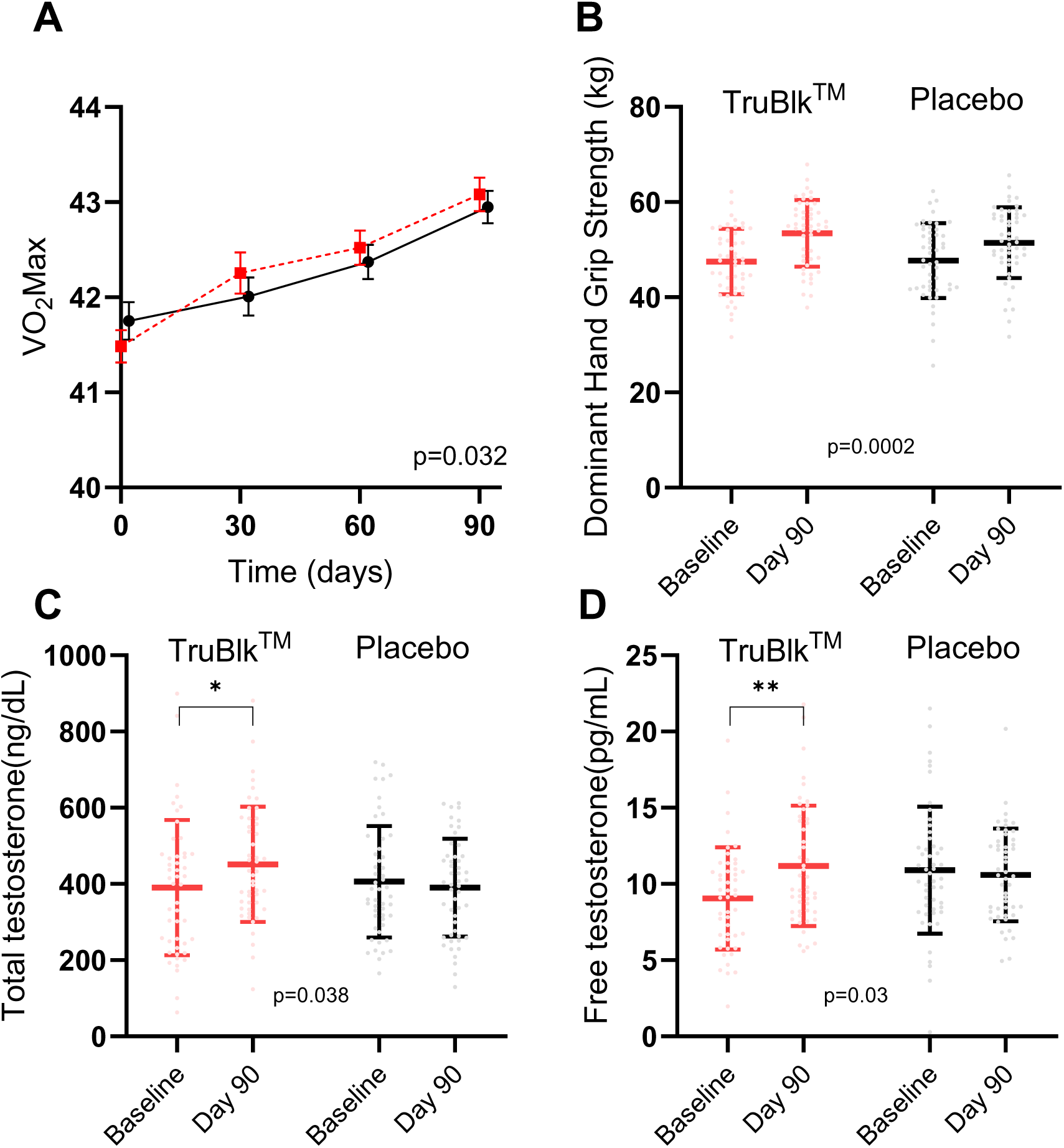
Cardiorespiratory fitness, grip strength and androgen status. (A) VO₂max, (B) dominant handgrip strength, (C) total testosterone and (D) free testosterone in participants receiving TruBlk™ (red, n = 47–49) or placebo (black, n = 50). VO₂max is shown across all four timepoints as mean ± SEM; remaining panels show baseline and day 90 values as mean ± SD, with individual participant values overlaid. Asterisks denote within-group change from baseline (*p < 0.05, **p < 0.01); P values denote the between-group difference in change from baseline.

#### Handgrip Strength

A more pronounced effect was seen for dominant handgrip strength. Groups were closely matched at baseline (TruBlk™ 47.51 ± 6.86 kg; placebo 47.70 ± 7.84 kg; p = 0.90), and grip strength increased significantly in both arms over the 90-day period (both p < 0.0001). However, the magnitude of improvement differed between groups: participants receiving TruBlk™ gained 5.94 ± 3.26 kg (+12.5%) compared with 3.76 ± 2.33 kg (+7.9%) in the placebo group, a between-group difference of 2.18 kg (95% CI 1.05 to 3.31) that represented a moderate effect (Cohen’s d = 0.77, p = 0.0002) (Figure 3B).

#### Testosterone

Total and free testosterone were assessed at baseline and day 90. Total testosterone was comparable between groups at baseline (TruBlk™ 390.7 ± 177.5 ng/dL; placebo 405.9 ± 146.2 ng/dL; p = 0.65), whereas free testosterone was modestly lower in the TruBlk™ group (9.05 ± 3.36 vs 10.91 ± 4.17 pg/mL; p = 0.018). Over the 90-day period, both measures increased in the TruBlk™ group and were unchanged in placebo. Total testosterone rose by 60.9 ± 172.8 ng/dL (+15.6%; within-group p = 0.019) in the TruBlk™ group versus −15.6 ± 185.9 ng/dL in placebo (p = 0.56), a between-group difference of 76.5 ng/dL (95% CI 4.4 to 148.5; Cohen’s d = 0.43, p = 0.038). Free testosterone rose by 1.97 ± 4.74 pg/mL (+21.8%; within-group p = 0.007) versus −0.32 ± 5.41 pg/mL in placebo (p = 0.68), a between-group difference of 2.28 pg/mL (95% CI 0.23 to 4.34; Cohen’s d = 0.45, p = 0.030). Mean values in both groups remained within the normal physiological reference range throughout (Figure 3C-D).

#### Muscle Damage Markers

Muscle damage markers were assessed at baseline and day 90. As these variables were markedly skewed and non-normally distributed, they are presented as medians with interquartile ranges and analysed non-parametrically, with Cliff’s delta used as the effect size. C-reactive protein was comparable between groups at baseline (p = 0.79) and remained unchanged in both arms over the study period, with no between-group difference in change (median +12.0% in the TruBlk™ group vs +14.0% in placebo; p = 0.95, Cliff’s δ = 0.01, Figure 4A). By contrast, both creatine kinase and lactate dehydrogenase declined more markedly in the TruBlk™ group. Creatine kinase fell by a median of 78.1 U/L (−33.6%) in the TruBlk™ group, from 226.2 to 153.0 U/L (within-group p < 0.001), compared with a fall of 9.8 U/L (−6.9%) in placebo, from 161.6 to 154.9 U/L (p = 0.47), with a significant between-group difference of small effect size (p = 0.030, Cliff’s δ = −0.26, Figure 4B). Lactate dehydrogenase followed the same pattern, falling by a median of 51.3 U/L (−17.8%) in the TruBlk™ group, from 281.8 to 219.3 U/L (p < 0.001), versus 10.3 U/L (−4.2%) in placebo, from 249.3 to 227.3 U/L (p = 0.047), again favouring the TruBlk™ group (p = 0.039, Cliff’s δ = −0.24, Figure 4C).

**Figure 4.**
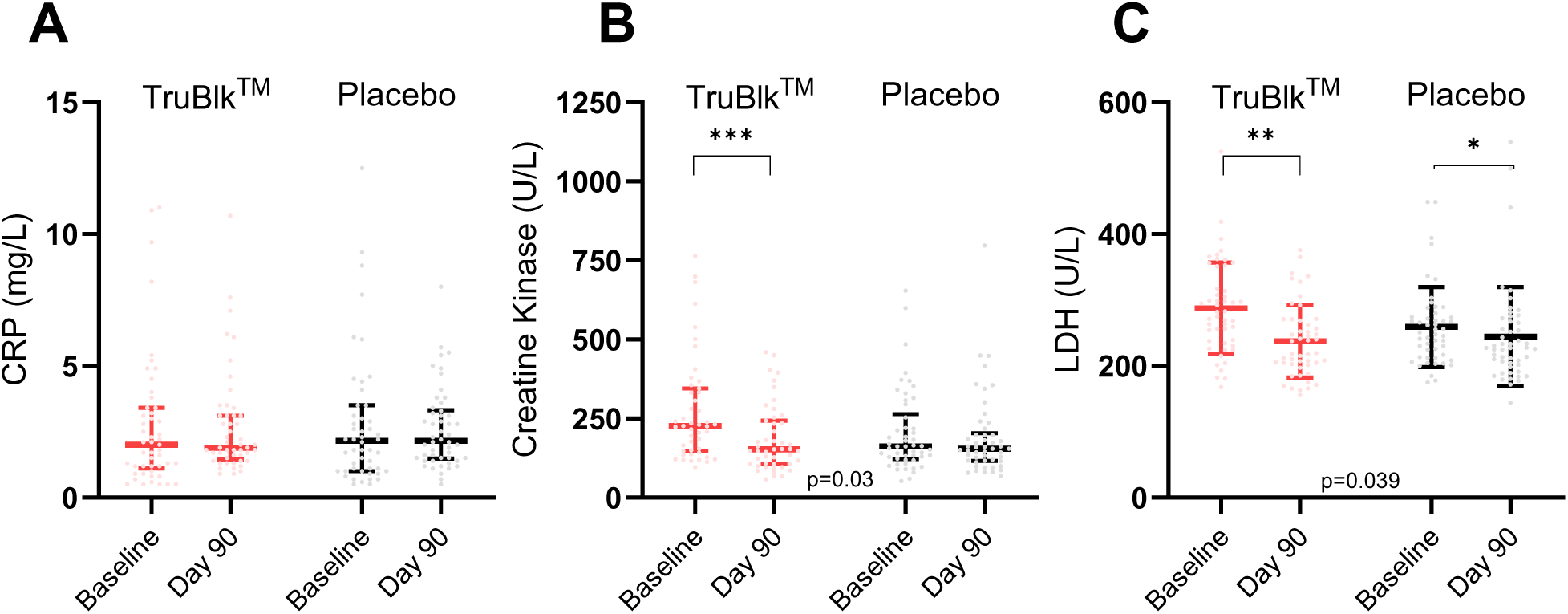
Muscle damage markers at baseline and day 90. **(A)** C-reactive protein (CRP), **(B)** creatine kinase (CK) and **(C)** lactate dehydrogenase (LDH) in participants receiving TruBlk™ (red, n = 49) or placebo (black, n = 50). Boxes show median and interquartile range with individual participant values overlaid. Asterisks denote within-group change from baseline (*p < 0.05, **p < 0.01, ***p < 0.0001); P values denote the between-group difference in change from baseline.

#### Global Assessments

At Day 90, 80% of TruBlk™ participants rated their improvement as “good” or “excellent” on the participant global assessment, compared with 60% in the placebo group. Investigator global assessment ratings were consistent with participant self-reports. The number needed to treat for a clinician-rated “good” or “excellent” global improvement response was 5.

### 3.4 Safety

TruBlk™ was well tolerated throughout the 90-day intervention period. No adverse events were recorded in the active treatment group. One mild, self-limiting gastrointestinal event was reported in the placebo group. Vital signs and anthropometric measurements remained stable in both groups across all timepoints. No serious adverse events occurred in either group

## 4. Discussion

This randomised, double-blind, placebo-controlled, multicenter trial is the largest evaluation of standardised Shilajit resin in resistance-trained males to date. Over 90 days, TruBlk™ Shilajit resin (250 mg twice daily; ≥60% fulvic acid, ≥10% shilarathenes™) produced significant improvements across all four primary endpoints versus placebo: maximal strength (1RM leg press), muscle endurance, rating of perceived exertion, and delayed-onset muscle soreness; with effect sizes in the small-to-moderate range. These gains were consistent across secondary endpoints, where dominant handgrip strength showed the largest between-group effect and the active group also demonstrated favourable shifts in muscle-damage markers, testosterone, and aerobic capacity. With 99% retention and no adverse events in the active group, the trial establishes a coherent efficacy–safety profile spanning strength, endurance, recovery, and hormonal domains.

These findings consolidate a fragmented evidence base. The testosterone increases align with Pandit et al. (2016), who reported comparable gains at an identical dose and duration, though in an older cohort (45–55 years); our data suggest the effect may be reproducible in younger trained males with baseline values already in the normal range [10]. The strength signal echoes Keller et al. (2019), whose benefits on fatigue-induced strength loss and hydroxyproline emerged only in the 500 mg/day group, supporting a dose-dependent threshold matching the total daily dose used here [9]. The CK and LDH reductions address a gap left by Das et al. (2016), who found upregulation of muscle-repair genes without corresponding marker changes, plausibly because that cohort lacked an exercise stimulus [20], whereas our requirement of regular training may have captured the functional consequence of those transcriptional changes. Finally, the results scale up the 28-day open-label TruBlk™ pilot within an adequately powered, blinded design [21]. A recurring constraint across this prior literature is the substantial variability in bioactive content and purity between most commercial Shilajit preparations, which plausibly explains inconsistent or dose-threshold-dependent effects — though this variability is not universal, as standardized, rigorously tested preparations from select suppliers demonstrate that consistent, verified bioactive content is achievable. The HPLC-validated standardisation guaranteeing ≥60% fulvic acid and ≥10% shilarathenes™ is therefore not merely a quality-control measure but a prerequisite for delivering bioactives at the concentrations required to elicit reproducible clinical effects, and is likely central to the consistency of the responses observed in this trial.

The magnitude of these effects is practically meaningful for a trained population. Resistance-trained individuals typically exhibit diminishing returns as their training experience accumulates, so a between-group strength and endurance advantage of the size seen here, achieved over established habitual training, represents a meaningful increment. The moderate effect sizes observed for the strength outcomes (d=0.60–0.77) are consistent with the magnitude expected of a nutritional supplement rather than a pharmacological agent. Notably, the consistency of direction across independent strength, endurance, perceptual and biochemical outcomes lends confidence that these represent genuine physiological effects rather than chance findings. This positions standardised Shilajit resin as a credible candidate among evidence-based ergogenic aids, while offering a complementary hormonal and recovery profile. The between-group testosterone difference reflected both a rise in the active arm and a small concurrent decline in the placebo group (−2.9% free, −3.8% total). The latter is of uncertain significance and falls within the range of ordinary biological and assay variation. Notably, values in the active group remained within the normal reference range throughout, consistent with a regulated rather than supraphysiological effect.

The biological rationale rests on complementary actions of the standardised bioactive fractions, though it should be emphasised that these mechanisms are inferred from preclinical and in vitro work rather than demonstrated in this cohort. Fulvic acid and dibenzo-α-pyrones have been proposed to act as electron shuttles in the mitochondrial electron transport chain, potentially enhancing ATP turnover, and preclinical work suggests activation of biogenesis through the PGC-1α/SIRT1 axis [14, 26]. Should such mechanisms operate in humans, they could plausibly underlie the endurance and RPE benefits observed, given the established role of mitochondrial content in fatigue resistance [27, 28]. Urolithin metabolites have been reported to support mitochondrial quality control via mitophagy [29], which may in principle help sustain respiratory capacity across repeated bouts of exercise.

The phenolic acid and flavonoid constituents of shilarathenes™ offer a plausible, though untested, explanation for the recovery and vascular findings [8]. Phenolic compounds are dietary antioxidants that have been shown to counteract exercise-induced oxidative damage and suppress inflammation [17, 30, 31], a mechanism that would be congruent with the CK and LDH reductions observed here. Flavonoids have been reported to increase endothelial eNOS activity and nitric oxide bioavailability [32], which could enhance blood flow and oxygen delivery to working muscle [33]; this represents one possible contributor to the modest VO₂max gain, which was of small magnitude and should be regarded as suggestive rather than definitive.

The increases in free and total testosterone are consistent in direction with the direct testicular actions reported by Pandit et al. (2016), which included upregulation of steroidogenic enzymes, aromatase inhibition, and improved Leydig cell responsiveness without hypothalamic-pituitary suppression [10]. The maintenance of values within the physiological range in this trial is at least compatible with such a regulated, non-suppressive mechanism, although the present study measured neither the enzymes nor the gonadotrophins required to test this directly. These hormonal changes were of modest magnitude and represent exploratory secondary outcomes; confirmation in a trial powered for endocrine endpoints would be required before firm conclusions are drawn.

These mechanisms therefore remain inferential and would benefit from direct interrogation in future work. As a starting point, high-resolution respirometry in muscle biopsies could quantify mitochondrial ATP flux. Building on Das et al. (2016), exercise-stimulated transcriptomic profiling would then map PGC-1α/SIRT1 and mitophagy activation [20]. Additionally, serial measurement of oxidative-stress and inflammatory mediators such as IL-6, TNF-α, and antioxidant enzymes would help test the antioxidant hypothesis. A dedicated endocrine study measuring SHBG, LH, FSH, and the testosterone-to-oestradiol ratio could help disentangle the relative contributions of steroidogenesis and aromatase inhibition. Finally, fraction-isolation studies comparing whole TruBlk™ against its purified constituents would establish which components drive each effect, and whether the proposed synergy is real.

Several limitations nonetheless temper interpretation. First, the cohort was restricted to healthy males aged 21–50, so the findings cannot readily be extrapolated to females, older adults, untrained individuals, or clinical populations. In addition, diet and training were habitual and self-regulated rather than controlled and may therefore have acted as a confounder. The secondary endpoints were exploratory and not individually powered, and the study was not designed to correct for multiplicity across them; the biochemical and hormonal findings should therefore be regarded as hypothesis-generating. Finally, the invoked mechanisms are extrapolated from preclinical literature, since no tissue-level measurements were undertaken in this cohort.

In conclusion, ninety days of standardised TruBlk™ Shilajit resin produced consistent improvements in strength, endurance, perceived exertion, and recovery in resistance-trained males, with favourable muscle-damage and testosterone shifts and an excellent safety profile. These results provide the first adequately powered, multicenter, placebo-controlled evidence that standardised full-spectrum Shilajit resin functions as a safe, effective ergogenic aid in a trained population, and establish a strong rationale for the mechanistic and longer-term studies outlined above.

## Data Availability

All data produced in the present study are available upon reasonable request to the authors

